# The elusive abscopal effect after radiotherapy: A systematic review and meta-analysis of the published literature

**DOI:** 10.1101/2025.04.30.25326672

**Authors:** Destie Provenzano, Murray Loew, Sharad Goyal, Yuan James Rao

## Abstract

**Background:** The abscopal effect is a phenomenon where a tumor that is not the intended site of treatment also shrinks in response to therapy; and is one of the most coveted and rare effects of cancer therapies. Previous attempts to summarize literature on the abscopal effect have focused on specific disease sites or been done on a limited basis. We performed a comprehensive review of every publication and case report regarding the abscopal effect induced by radiotherapy from the 1950s to February 2023 to identify potential trends and patterns in its induction based on specific disease/ treatment site, radiation dose, and other factors.

**Methods:** Literature that described the abscopal effect was identified through search of online databases: Pubmed, Medline, and Google Scholar for all published articles from 1953 leading to February of 2023 that included the terms “Abscopal Effect” and “Radiation” or “Radiotherapy.” Demographics (patient and tumor characteristics), radiation and abscopal effect analysis, outcome and treatment analysis, and risk of bias were reported and summarized.

**Results:** A final total of 92 papers corresponding to 99 separate lesions or cases were included in the meta-analysis. The most common site of induction of the abscopal effect and of the target of the abscopal effect was the lung. One manuscript reported a nonstandard treatment protocol, the remainder of cases received radiation within reasonable treatment guidelines. The majority of manuscripts included most important information (cancer site, number of prior treatments, histopathology, outcome information, and dose).

**Conclusion:** Additional research is urgently needed to understand how and why some treatment regimes are able to induce the abscopal effect. For the time being, this remains a rare phenomenon.

## I. Introduction

The abscopal effect is one of the most coveted and rare effects of cancer therapies. The National Cancer Institute (NCI) defines the abscopal effect as a phenomenon where a tumor that is not the intended site of treatment also shrinks in response to therapy. [1] The abscopal effect was first observed by Mole in the 1950s when radiotherapy induced a clinical response at distant tumor site than the target. [2] The abscopal effect is considered to have occurred when reduction of 30% of a tumor distant from the site of treatment is observed. [3] Cancer therapies such as radiation therapy seek to reduce tumor mass and induce DNA damage through double-stranded breaks and the generation of reactive oxygen species, which can damage surrounding tissue. [4] The induction of the abscopal effect increases the impact of targeted radiation therapy without increasing toxicity to normal tissue, making it a desirable effect for radiation therapy. [5] Additionally, attempts to induce the abscopal effect can be seen as a potential last resort for patients with advanced metastatic cancer such as lung cancer, who traditionally do not have many options for treatment. [6] The abscopal effect is unfortunately incredibly rare, with only a handful of cases reported since it was first described.

Various studies have attempted to induce the abscopal effect through radiation therapy without additional treatments with unpredictable results. [7,8] However immunotherapies used in combination with radiation therapy have shown to induce the abscopal effect. [9, 10, 11] Immunotherapies involving immune checkpoint inhibitors (ICIs): CTLA-4, PD-1 and PD-L1, have previously been shown to work in conjunction with radiotherapy to induce the abscopal effect. [12]

Previous attempts to summarize literature for the abscopal effect have been done on a disease site specific or otherwise limited basis. [13, 14, 15] We performed a comprehensive review of every publication and case report regarding the abscopal effect from the 1950s to February 2023 to identify potential trends and patterns in its induction based on specific disease/ treatment site, radiation dose, and other factors. For our final analysis we sought to include only studies that reviewed the abscopal effect due to radiotherapy or a combination of radiotherapy and immunotherapy.

## II. Methods

### A. Manuscript Identification

Literature that described the abscopal effect was identified through search of online databases: Pubmed, Medline, and Google Scholar. All published articles from 1953 leading to February of 2023 were considered for the purpose of this review. Inclusion criteria for this study included articles that included the terms “Abscopal Effect” and “Radiation” or “Radiotherapy.” Studies were excluded if they didn’t report on abscopal effort due to radiotherapy (for example if instead they saw the abscopal effect used for HIFU treatment), if the article was not in English, if the article reported on the abscopal effect in a non-human (such as mouse) model, or if the article contained insufficient radiotherapy data.

Operationally, the radiation induced abscopal effect was defined as any case of malignancy in which 1) There were at least two separate anatomical sites of cancer, of which at least one of them is measurable (index lesion) 2) ionizing radiation therapy was administered to at least one site, but not to the index lesion; and 3) the index lesion was reported to decrease in size without known exposure to radiotherapy. Articles describing clinical scenarios not meeting the above criteria were excluded. Articles describing treatments of radiation in combination with surgery or systemic therapy were allowed. Allowed methods of ionizing radiation included x-ray photons, electrons, heavy particles such as protons or carbon ions, brachytherapy, or radio-isotopes.

Articles were reviewed twice by two of the authors: a cancer researcher (Graduate student, DP) and radiation oncologist (Board-certified, YR) to ensure integrity of final summarized results. Articles were reviewed for criteria summarized in Table 1. Full Inclusion and Exclusion Criteria is listed in Table 1. A table of all included articles and abstracted data is included in the Supplemental Appendix (Table S1). PRISMA Flowchart and Checklist are detailed in the Supplemental Appendix (Figure S1, Table S2).

**Table 1:** Summary of inclusion criteria, exclusion criteria, and topics evaluated within selected articles.

| Topic | Terms |
| --- | --- |
| Inclusion Criteria | Terms "Abscopal Effect" from 1953 to February 2023 |
| Exclusion Criteria | Reported on Abscopal Effect due to causes other than Radiotherapy (Such as HIFU) |
|  | Article not in English |
|  | Insufficient Radiotherapy Data |
| Topics Evaluated | Primary Tumor Site |
|  | Site of abscopal effect |
|  | Previous treatment |
|  | Site of radiotherapy |
|  | Radiotherapy dose (free text) |
|  | Total Dose (Gy) |
|  | Fractions |

### B. Demographics (Patient and Tumor Characteristics)

Primary tumor site, site of abscopal effect, previous treatment, site of radiotherapy, RT type, use of immunotherapy, type of immunotherapy, and previous agents (Chemotherapy or targeted agents) are totaled and summarized. Distribution is reported for radiotherapy dose, total dose, fractions, dose, hypofractions, ultra-hypofractions, and BED10. Clinical measures of the size of RT treated tumor, size of index lesion prior to AE, Size of index lesion after AE, duration of AE response, and survival outcome of patient after RT are also reported.

For this study, the primary tumor site is defined as the organ in which the cancer originated. The index lesion site or abscopal lesion site is the organ in which the abscopal effect was observed. The treatment lesion site is the organ that was treated with radiotherapy. The abscopal or index site can be the same organ if there were multiple lesions and not all were treated.

### C. Radiation and Abscopal Effect Analysis

Total lesions identified across the articles for each primary tumor, treatment site, and final abscopal (index) site are summarized and depicted to demonstrate the distribution and connections between each. Total radiation dose is summarized and depicted by histogram to demonstrate the range of dosage previously seen to induce the abscopal effect. Total size and range of treatment site lesion, and abscopal (index) lesion before and after treatment is depicted by box plot.

All reported lesions from manuscripts marked for inclusion in this study were included in this analysis. If a manuscript reported multiple abscopal effects, all lesions were treated separately for the purpose of this analysis.

### D. Outcome and Treatment Analysis

Survival curves depicting overall survival and abscopal site local control (If the tumor remained regressed) were calculated using the Kaplan Meir method in Graphpad software. Survival curves depict the proportion of patients at each time point who remain in the group (alive or without local progression at the abscopal site).

Radiation Therapy Treatment (RT) type, use of immunotherapy, surgery, and previous agents (Chemotherapy or targeted agents) are tabulated and summarized.

All reported patients from manuscripts marked for inclusion in this study were included in this analysis.

### E. Risk of Bias

Risk of bias presented in the articles was evaluated for presence and quality of information by radiation oncologist for quality of the reported demographic and radiation therapy information, and for reliability of results. Articles were checked for inclusion of demographic criteria and tumor information. These included: Age, Sex, Histopathology, Number of prior lines of treatment, Size (RT treated, cm), Size (index before AE), Size (index after AE), Outcome, Correlative Biomarker (any measured by paper), Primary Tumor Site, Primary_Fixed, Site of abscopal effect, and Radiation Dose.

Manuscripts were also evaluated by radiation oncologist for if reported radiation dose and treatment protocol was within typical guidelines for treatment.

Prisma 2020 checklist for systematic review and meta analyses was used to ensure all potential biases and robustness of analysis evaluated. Full checklist is included in the supplement (FlowChart Figure S1, Checklist Table S2).

### F. Statistical Analysis

Statistical analyses included in this paper began with an exploratory data analysis of demographics (median, range, summary statistics, etc.). Histogram and box plot analyses were conducted to display continuous variables. Survival analysis was conducted in Graphpad to display outcome information.

## III. Results

### A. Manuscript Identification

A total of 180 papers were initially identified in literature search that referenced the abscopal effect from some form of treatment. Of these 88 were excluded due to use of a therapy different from radiotherapy (Cryoablation, HIFU, Y-90, etc.), reference to abscopal effect of occurring due to toxicity not therapy, language not in English, or if they contained insufficient data regarding the patient and course of action (no radiotherapy data, etc.). A final total of 92 papers corresponding to 99 separate lesions or cases were included in the meta-analysis. For the purposes of this manuscript, a case or lesion refers a primary, abscopal (index), or treated site of cancer.

### B. Demographics (Patient and Tumor Characteristics)

Demographics, the primary tumor, site of abscopal effect (index), and radiotherapy site listed in each of the studies was recorded and summarized. (Table 2) The tumor site published most frequently in literature regarding the abscopal effect was the lung (23 primary, 42 abscopal (index), 19 treatment site).

**Table 2:**
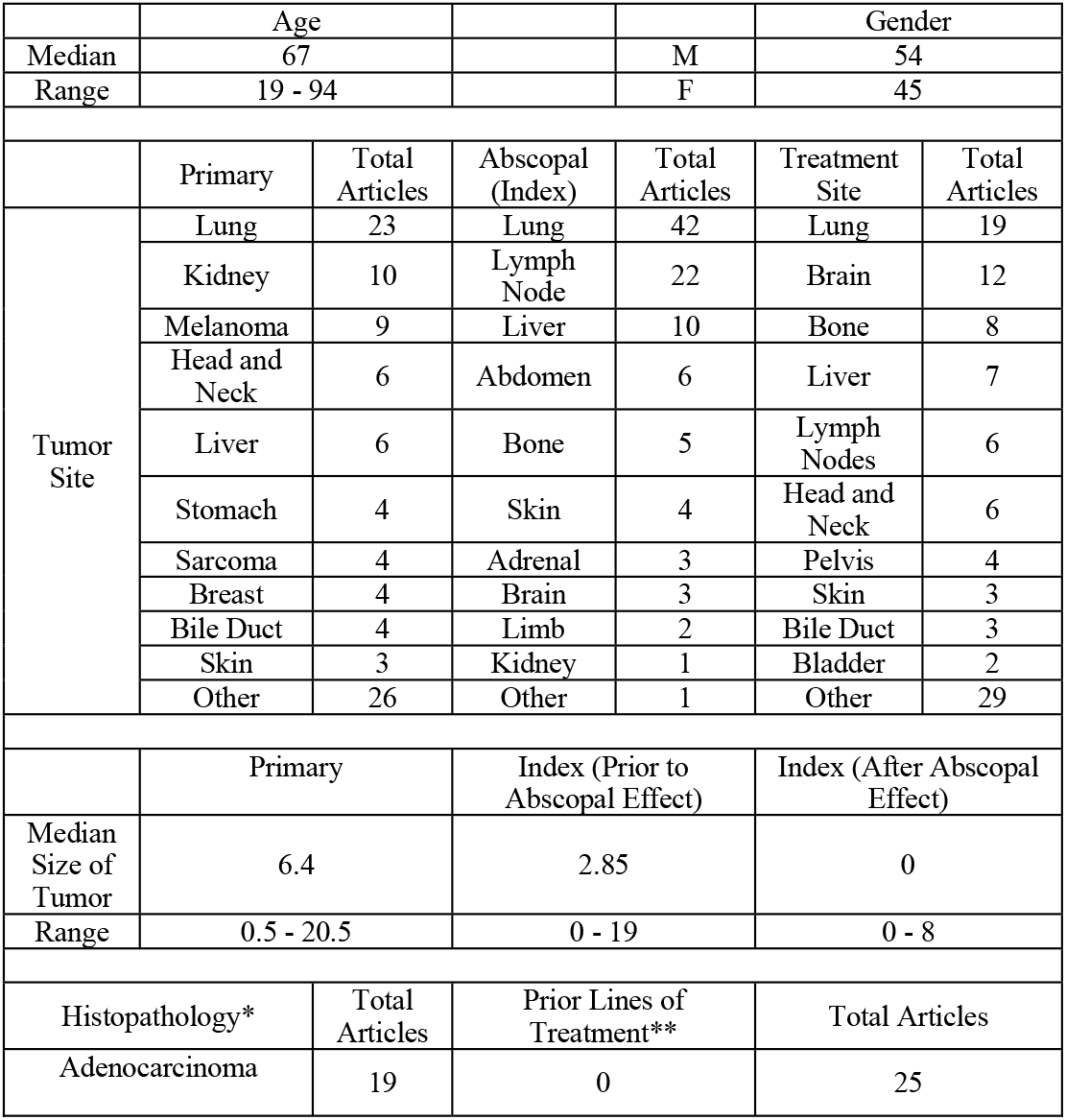
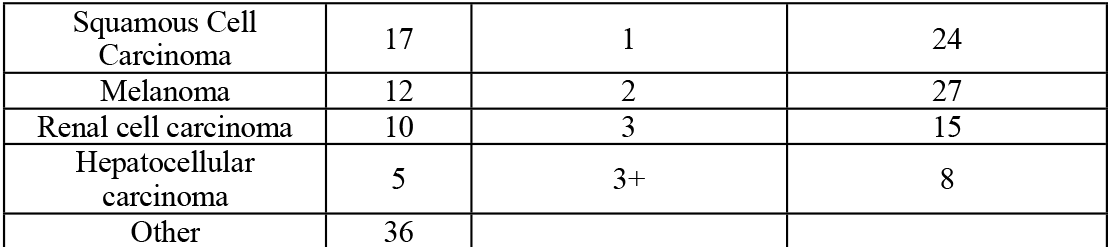
Summary of demographics and tumor information from analyzed manuscripts. *Histopathology is the histologic type of tumor reported by the pathologist based off of biopsy. **Prior lines of treatment are previous treatments administered in sequence prior to the abscopal effect. This may include prior surgery, radiation, chemotherapy or combinations of these. The 0, 1, 2, 3 and 3+ in the description describe the total number of prior treatments.

Most common primary site, treatment (radiotherapy) site, and abscopal (index) effect site pairings are depicted in Figure 1. Top 10 sites indicating primary tumor, treatment site, and abscopal effect are depicted, with remaining published tumor sites grouped into “other” category.

**Figure 1:**
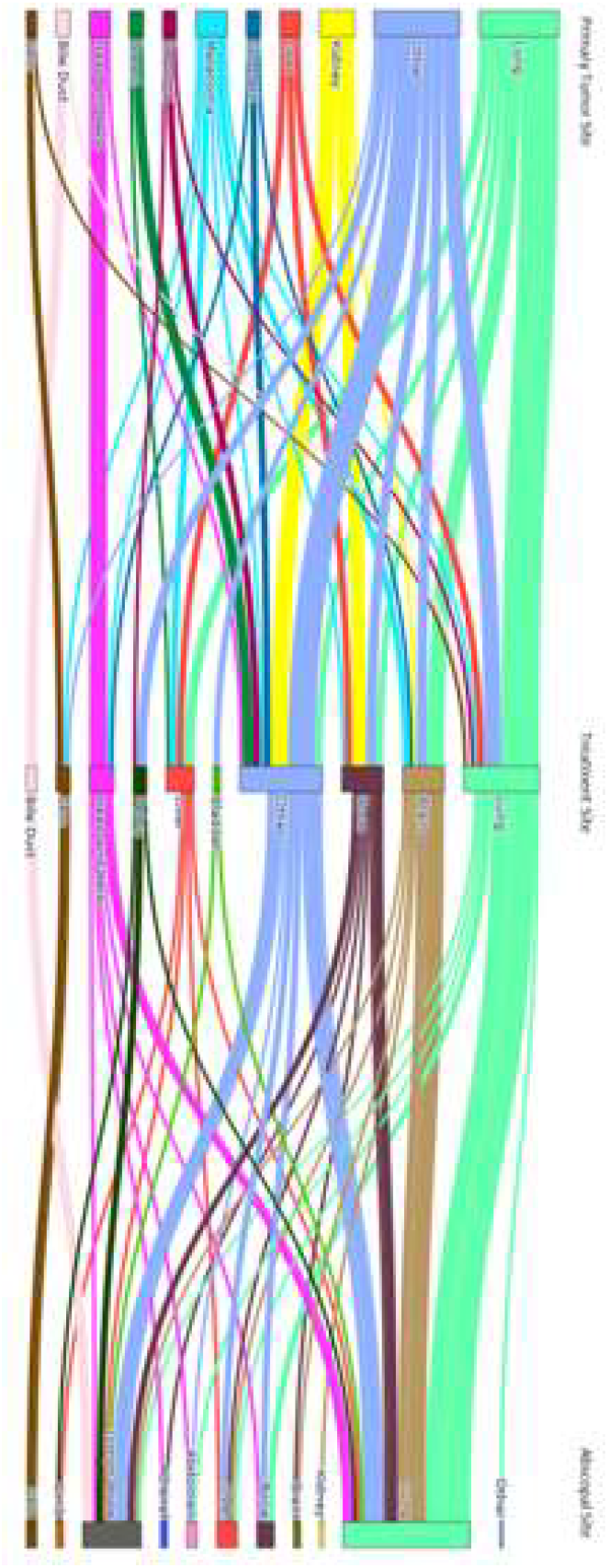
Primary tumor, abscopal (index) effect, and radiotherapy site listed in publications regarding the abscopal effect.

### C. Radiation and Abscopal Effect Analysis

Distribution for radiotherapy dose (Gy) and BED10 (gy) are depicted in Figure 2. Median Total Dose was 32 Gy, Fractions was 10, Dose/Fx was 3, Hypofractions (3+) was 1, Ultra-Hypofractions (5+) was 0, and BED10 was 46.80 Gy. Interquartile ranges for radiotherapy dose and BED10 were 30-48 Gy and 39-71.7 respectively.

**Figure 2:**
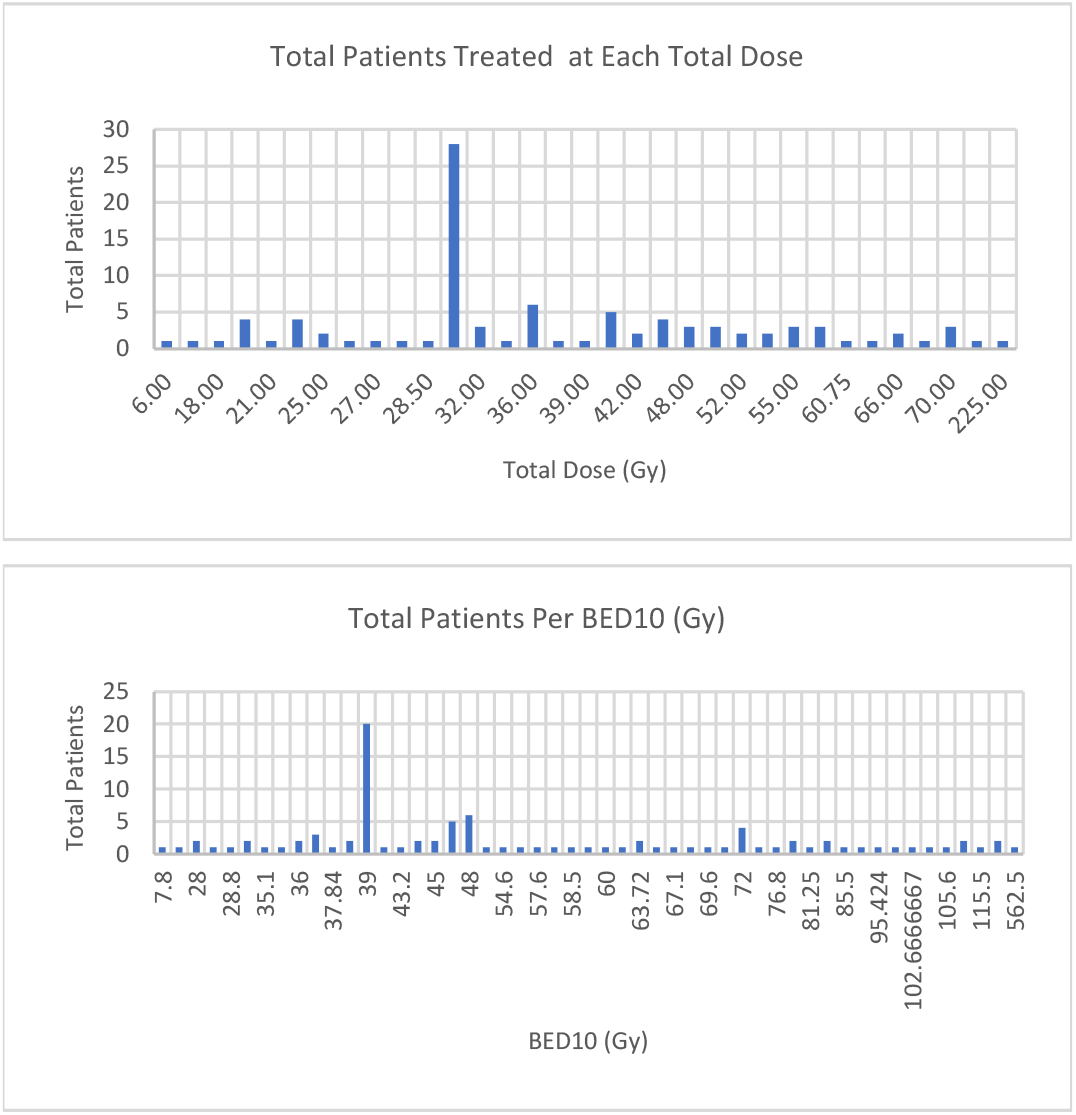
Total patients vs Radiation Dose (Gy), and total patients vs BED10 (Gy) are depicted in Figure 2.

### D. Outcome and Treatment Analysis

Boxplots depicting size of the index lesion before and after the abscopal effect and size of the tumor treated with radiation therapy are shown in cm in Figure 3.

**Figure 3:**
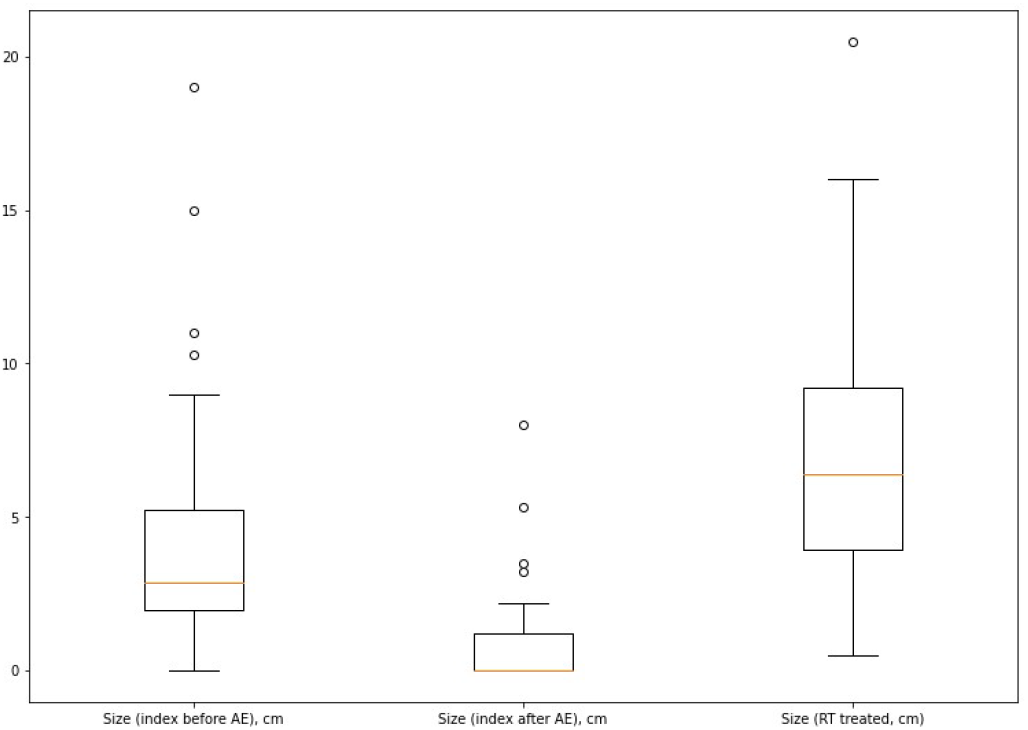
Size of treatment site (RT treated tumor) in cm, size of abscopal (index) lesion prior to abscopal effect in cm, Size of index lesion after abscopal effect in cm.

Survival curves for overall survival and abscopal site local control (did tumor remain regressed) are depicted in Figure 4.

**Figure 4:**
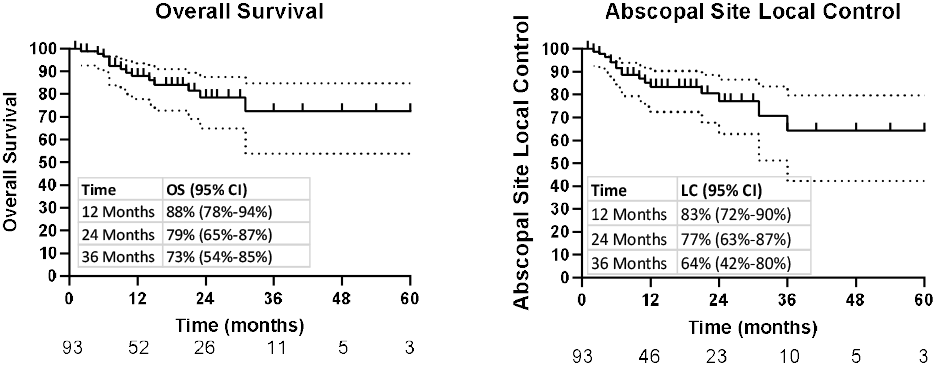
Survival curves depicting overall survival and abscopal site local control.

RT type, use of immunotherapy, surgery, and previous agents (Chemotherapy or targeted agents) are identified and summarized in Table 3.

**Table 3:** Prior and Current Treatments: Use of surgery (Y/N), Cytotoxic Chemotherapy (Y/N), Immunotherapy (Y/N), Previous Targeted Agents (Y/N), List of Targeted Agents, and Radiotherapy type.

|  | Yes | No | Not Reported |
| --- | --- | --- | --- |
| Surgery | 11 | 88 |  |
| Cytotoxic Chemotherapy | 62 | 33 | 4 |
| Immunotherapy | 56 | 39 | 4 |
| Previous Targeted Agents | 28 | 67 | 4 |
| If reported - Type of Targeted Agent |  | Total |  |
| Tyrosine Kinase Inhibitor |  | 11 |  |
| Hormone Therapy |  | 5 |  |
| EGFR Inhibitor |  | 4 |  |
| BRAF Inhibitor |  | 2 |  |
| Anti-HER2 Therapy |  | 1 |  |
| EZH2 inhibitor |  | 1 |  |
| VEGEF Inhibitor |  | 1 |  |
| BRAF and MEK inhibitors |  | 1 |  |
| Radiotherapy Type |  | Total |  |
| Brachytherapy |  | 3 |  |
| EBRT |  | 92 |  |
| EBRT plus Brachytherapy |  | 1 |  |
| TARE |  | 1 |  |
| Not Reported |  | 2 |  |

### E. Risk of Bias

One manuscript reported a nonstandard treatment protocol (225 Gy in 15 fractions). The remainder of cases received radiation within reasonable treatment guidelines.

Presence and quality of reported information and evidence present in the manuscripts is summarized in Table 4. The majority of manuscripts included most important information (cancer site, number of prior treatments, histopathology, outcome information, and dose). However many lesions reported by the manuscripts lacked information for various cancer site sizes (64 did not report RT treatment site size, 71 did not report abscopal (index) size before, and 73 did not report abscopal (index) size after) and for if a correlative biomarker was measured by the paper.

**Table 4:**
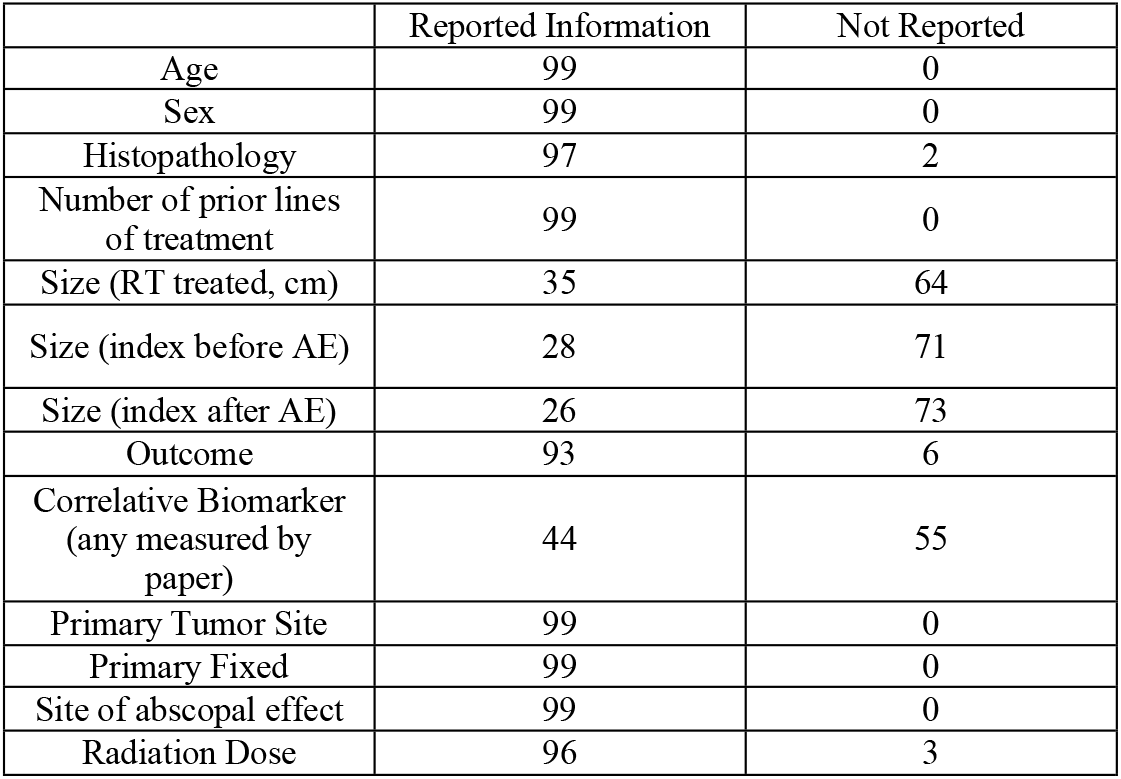
Inclusion of demographic and lesion (case) information for the 99 lesions reported across 92 manuscripts.

|  | Reported Information | Not Reported |
| --- | --- | --- |
| Age | 99 | 0 |
| Sex | 99 | 0 |
| Histopathology | 97 | 2 |
| Number of prior lines of treatment | 99 | 0 |
| Size (RT treated, cm) | 35 | 64 |
| Size (index before AE) | 28 | 71 |
| Size (index after AE) | 26 | 73 |
| Outcome | 93 | 6 |
| Correlative Biomarker (any measured by paper) | 44 | 55 |
| Primary Tumor Site | 99 | 0 |
| Primary Fixed | 99 | 0 |
| Site of abscopal effect | 99 | 0 |
| Radiation Dose | 96 | 3 |

## IV. Discussion

To our knowledge, no publications have reported more than two cases of the abscopal effect in a single report, and the vast majority (87) report a single case of the abscopal effect in a publication.

We observed that the abscopal effect was seen in a number of tumor types, most commonly lung, but also the kidney, liver, stomach, and other organs. The organ site in which the abscopal effect most often occurred was the lung, which was usually a different organ from which the radiotherapy was delivered.

62 patients received previous cytotoxic chemotherapy, and 56 received immunotherapy at the time of the abscopal effect. Surprisingly, it was found that the abscopal effect was sometimes reported in patients who were not heavily pre-treated and in fact 25 had the effect occur as part of the first course of treatment.

Regarding radiation, the majority of patients received hypofractionated treatment over a median of 32 Gy in 10 fractions, and the mean BED given in treatments was 46.80. Proton therapy, carbon ion therapy, and brachytherapy were also reported methods of inducing the abscopal effect. The effect was reported over a wide range of doses and BEDs, with the range from 6 to 225. The most common organ target that induced the abscopal effect was lung followed by brain.

This review was limited as manuscripts that used a therapy different from radiotherapy (Cryoablation, HIFU, Y-90, etc.), that were not written in English, or that referred to an “abscopal adverse effect” where injury occurred at a distant site instead were excluded. This review did not consider any abscopal effect outside of radiation, as such a more complete picture may be drawn by inclusion of other methodologies. The evidence portrayed in the identified manuscripts is limited in that it is self-reported and typically relies on retrospective case-reports. There persists little data as to how to induce the abscopal effect.

A radiation oncologist once said: “Everyone thinks that they might have seen the abscopal effect once, but nobody remembers how to find it.” We conclude that additional research is required to better understand and predict the abscopal effect after radiation therapy. Most importantly, we as a field must undertake the mission of determining the approximate baseline frequency of occurrence in cancer patients. If the incidence is determined to be extremely rare, we must critically examine whether this is a real phenomenon or possibly caused by other factors. Additionally, patients rarely may experience spontaneous tumor regression in the absence of a therapeutic intervention. Only once the baseline incidence is known will it be possible to design prospective trials to attempt to induce the abscopal effect.

The radiation induced abscopal effect remains a rare and fleeting phenomenon, and with no reliable method of inducing this effect in any human malignancy. In the current state of evidence, it is not advisable to administer radiotherapy seeking to induce the abscopal effect outside the context of a clinical trial designed specifically to measure this outcome. Additional preclinical and clinical research is needed to evaluate the radiation induced abscopal effect, and to develop treatment strategies that unleash its potential.

## V. Conclusion

This systematic review and meta-analysis analyzed total abscopal events published in literature from 1950 – 2023 and found that it is an extremely rare occurrence. Most abscopal effects occur in the lung, and many of these abscopal effects follow a primary and treatment site also located within the lung. Additional research is urgently needed to understand how and why some treatment regimes are able to induce the abscopal effect. For the time being, this remains a rare phenomenon.

## Data Availability

All data produced are available online at citations listed within the article.

## SUPPLEMENT

**Table S1:**
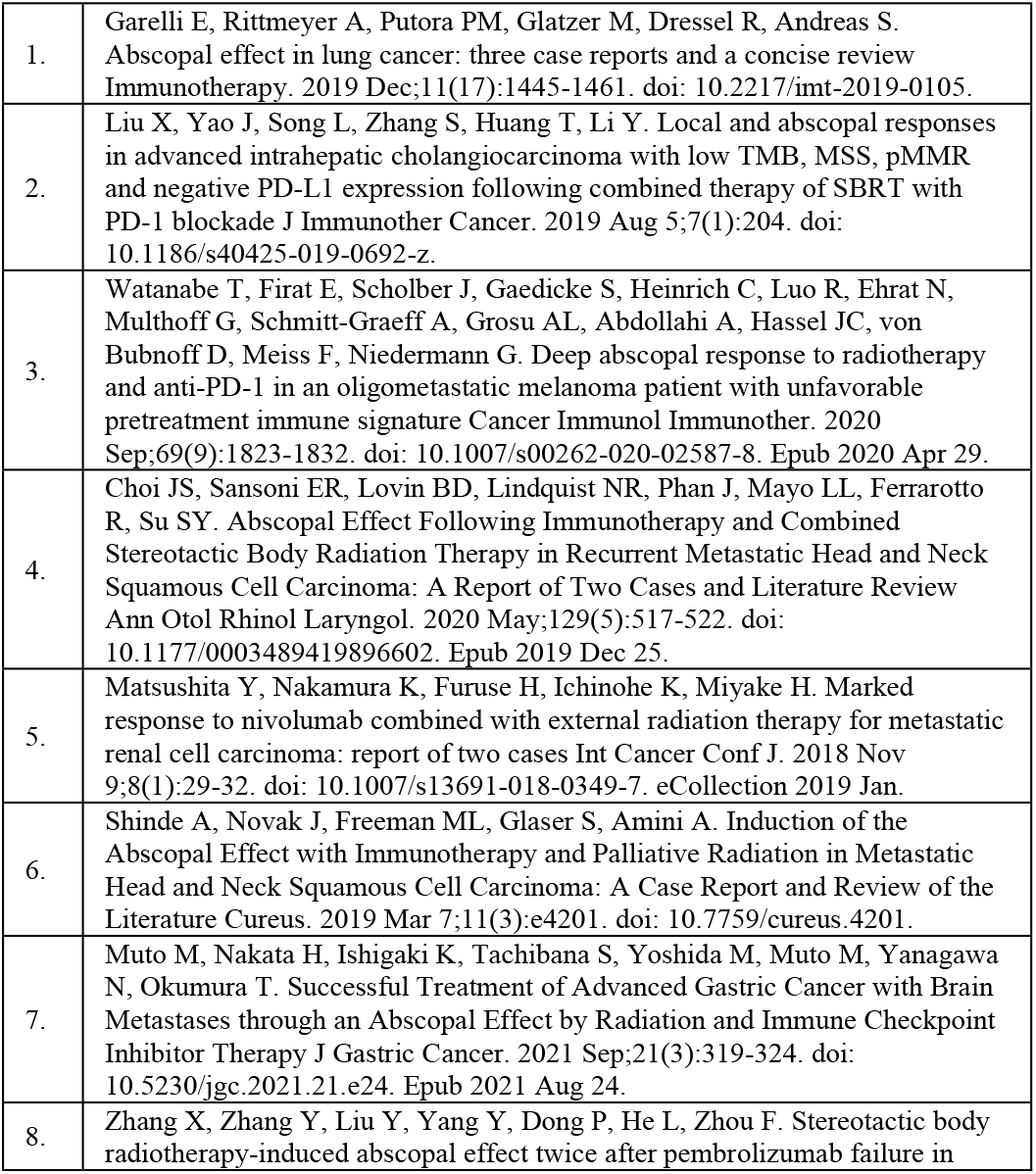
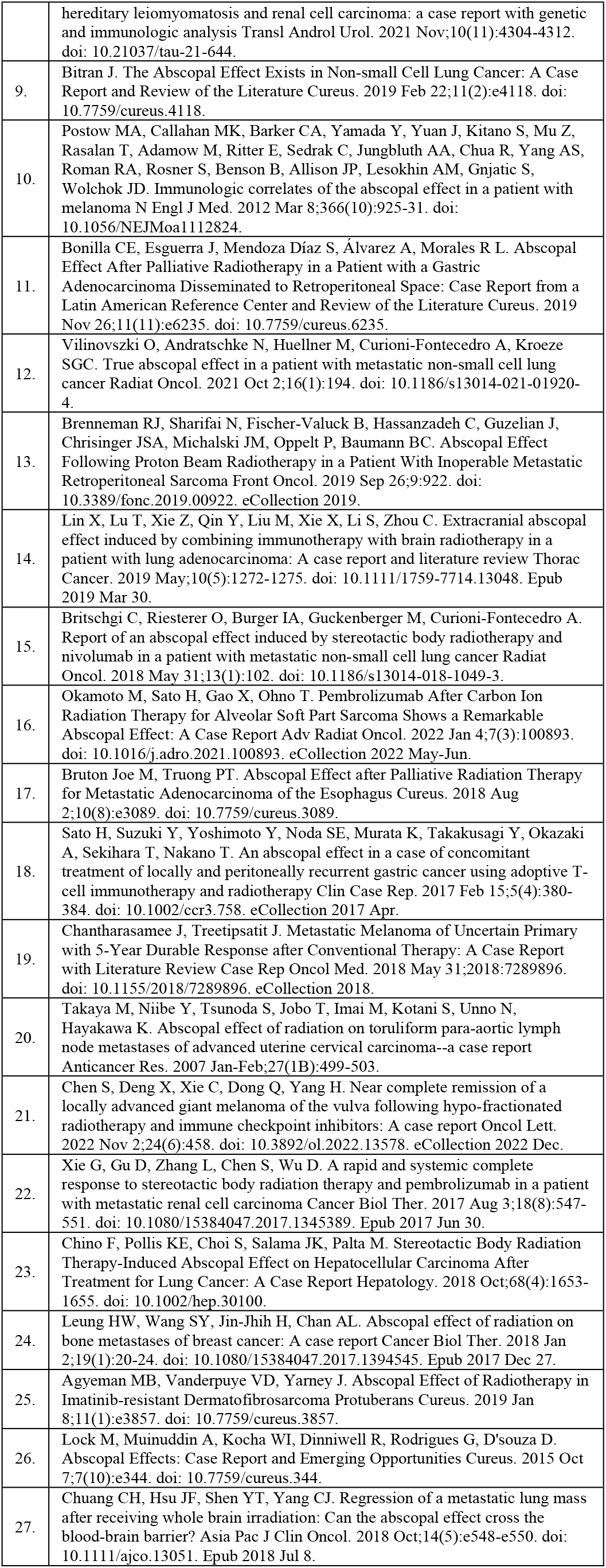
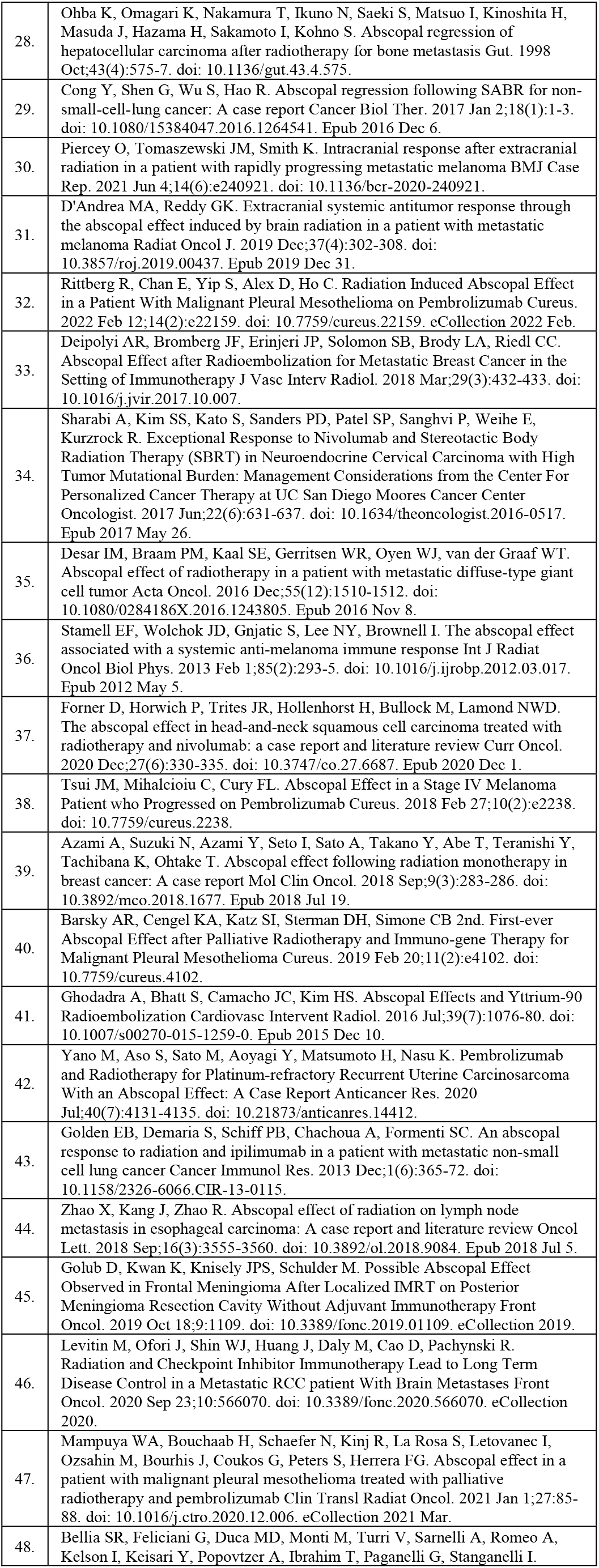
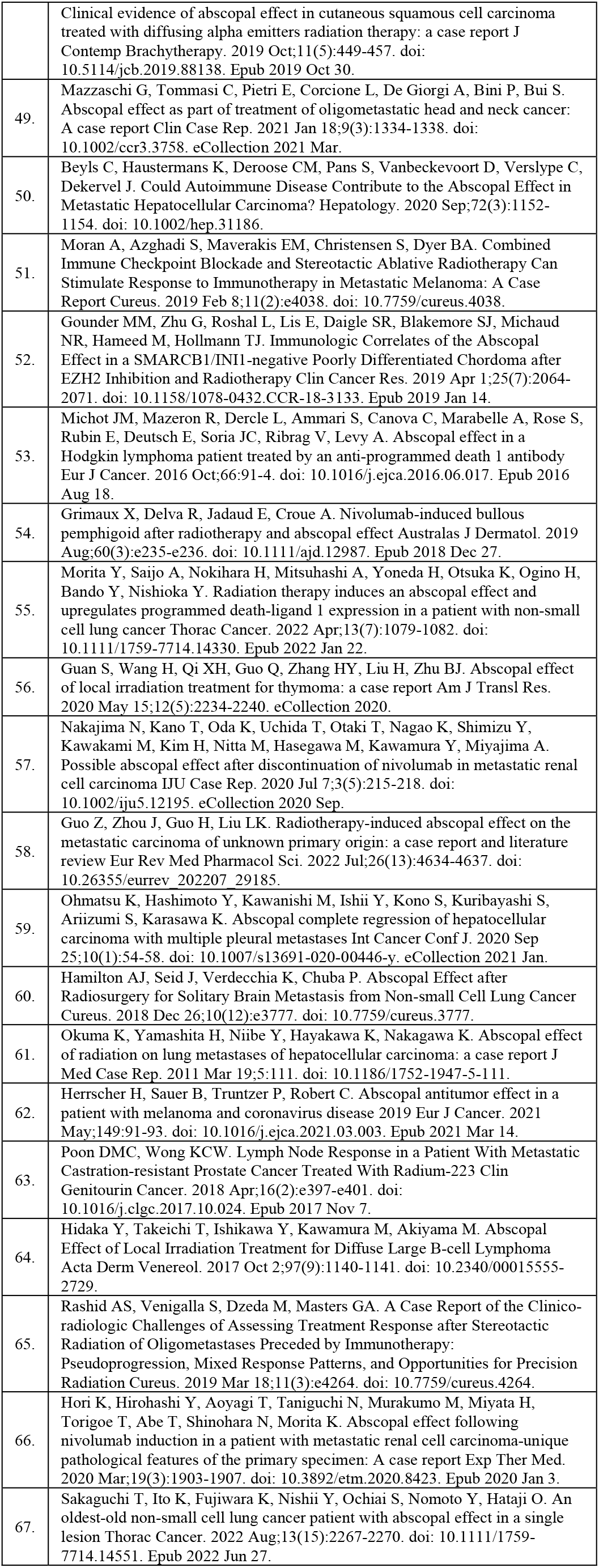
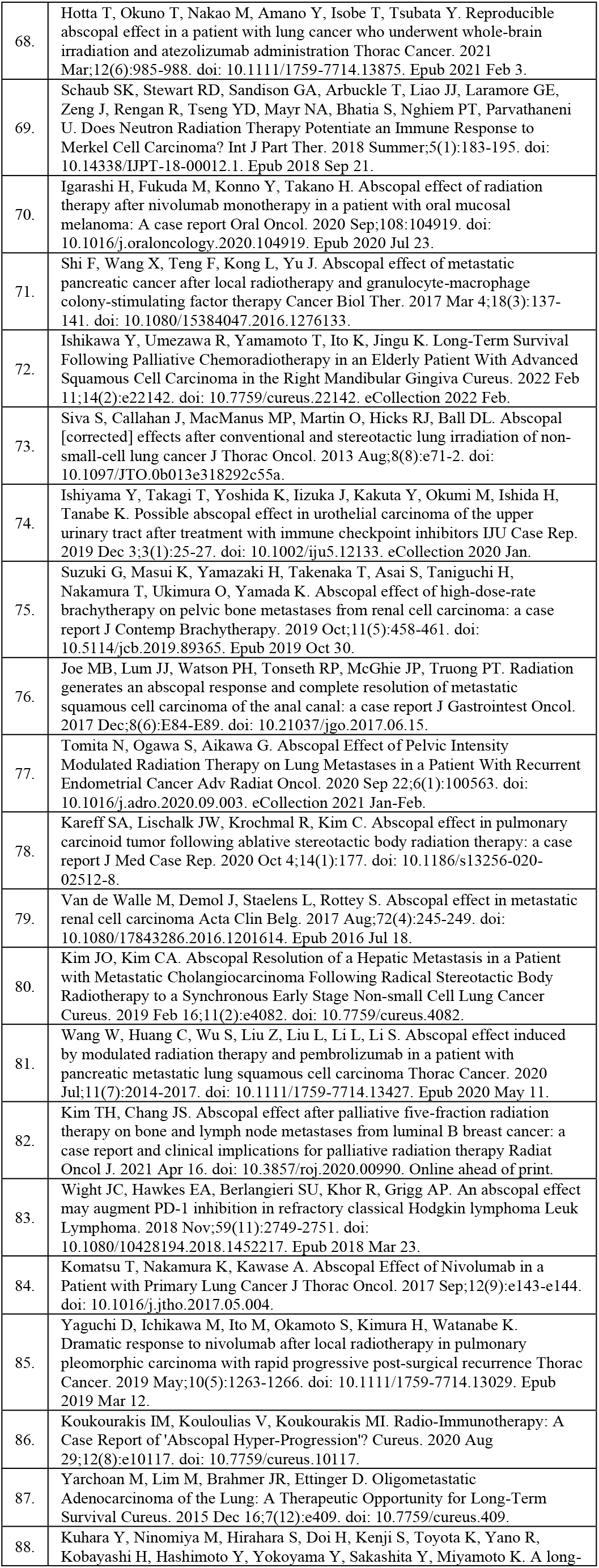
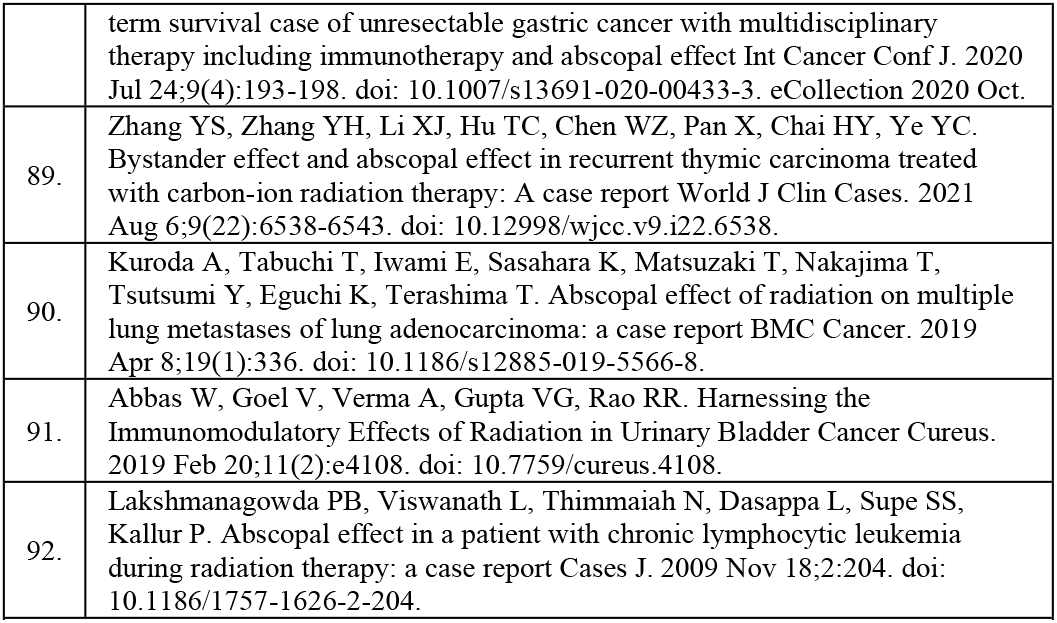
Full List of Articles Included in the Systematic Review and Meta Analysis.

**Figure S2:**
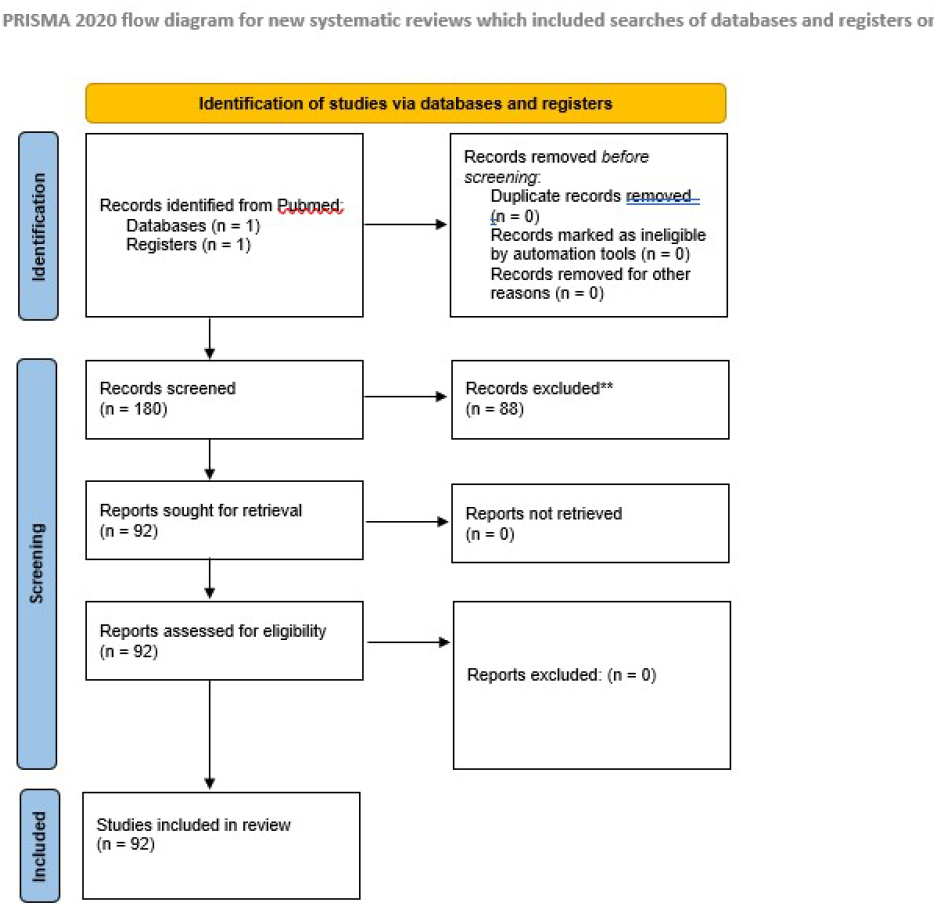
Full PRISMA 2020 FlowChart for Records screened and included in this systematic review and meta analysis. Source: Page MJ, et al. BMJ 2021;372:n71. doi: 10.1136/bmj.n71. This work is licensed under CC BY 4.0. To view a copy of this license, visit https://creativecommons.org/licenses/by/4.0/

**Table S3:**
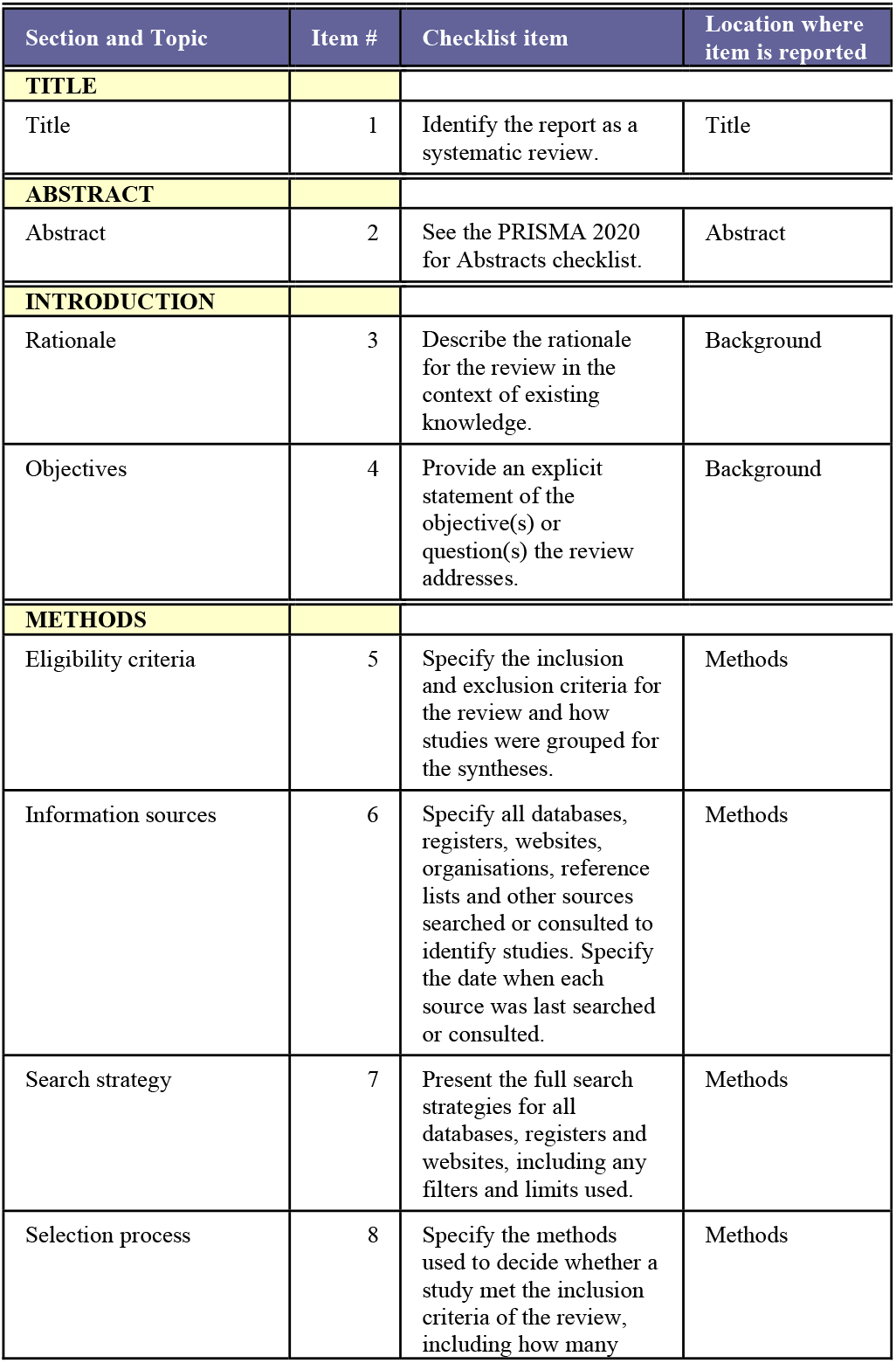
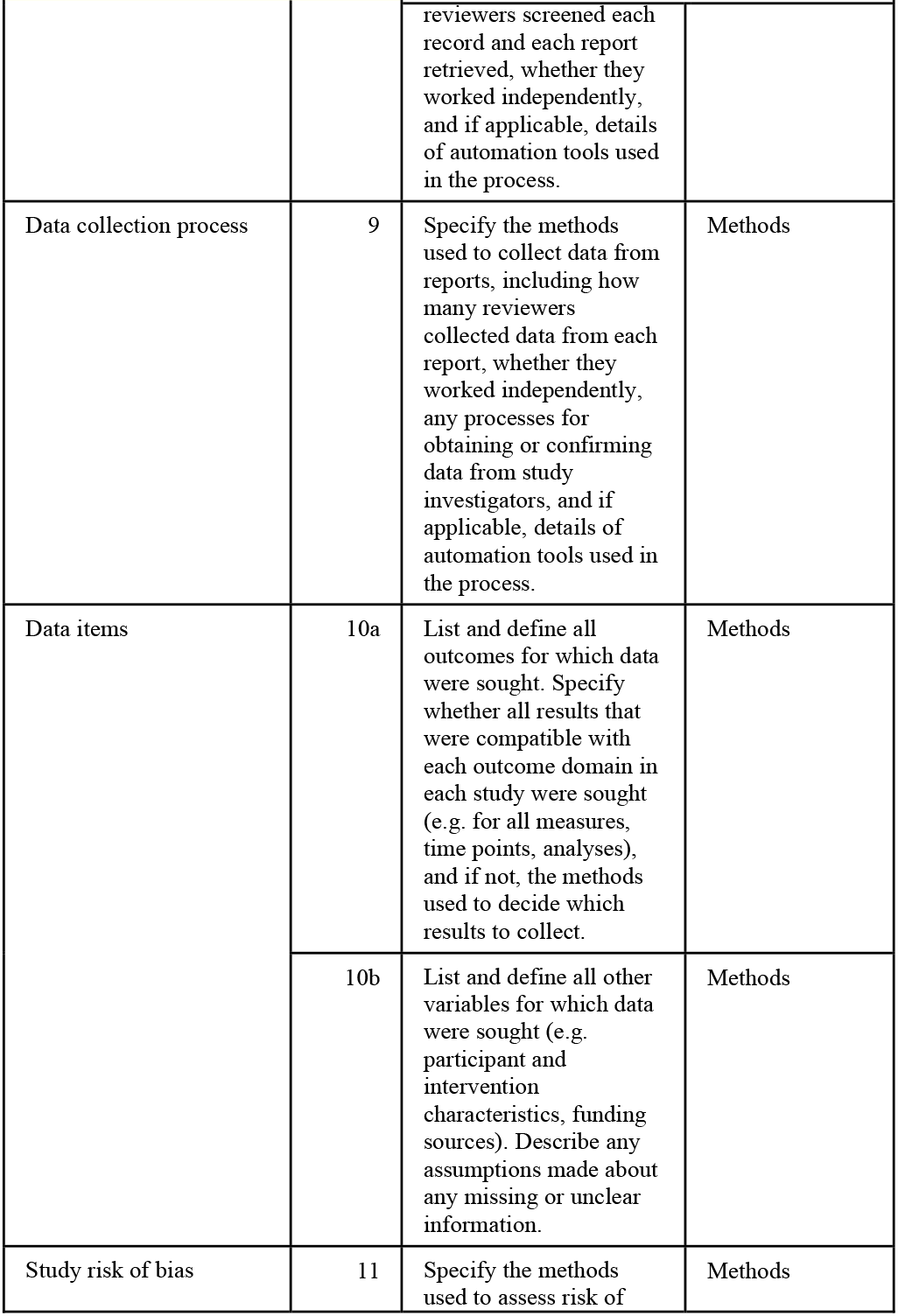
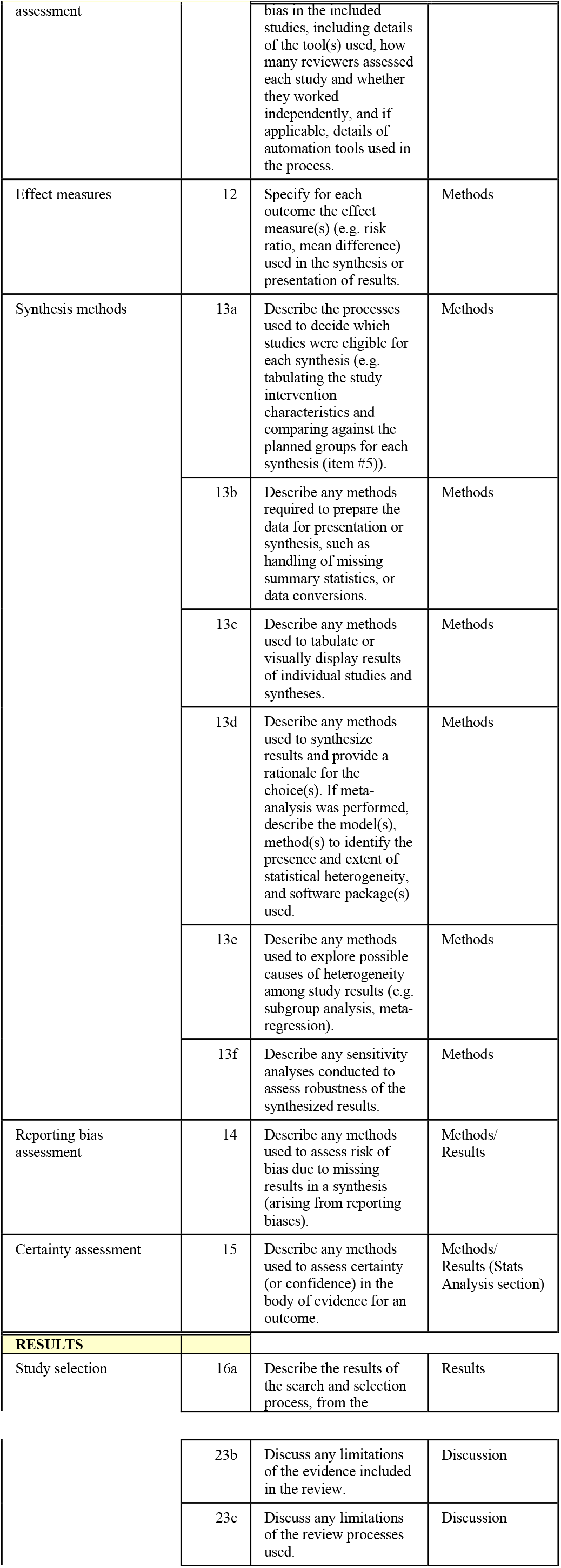
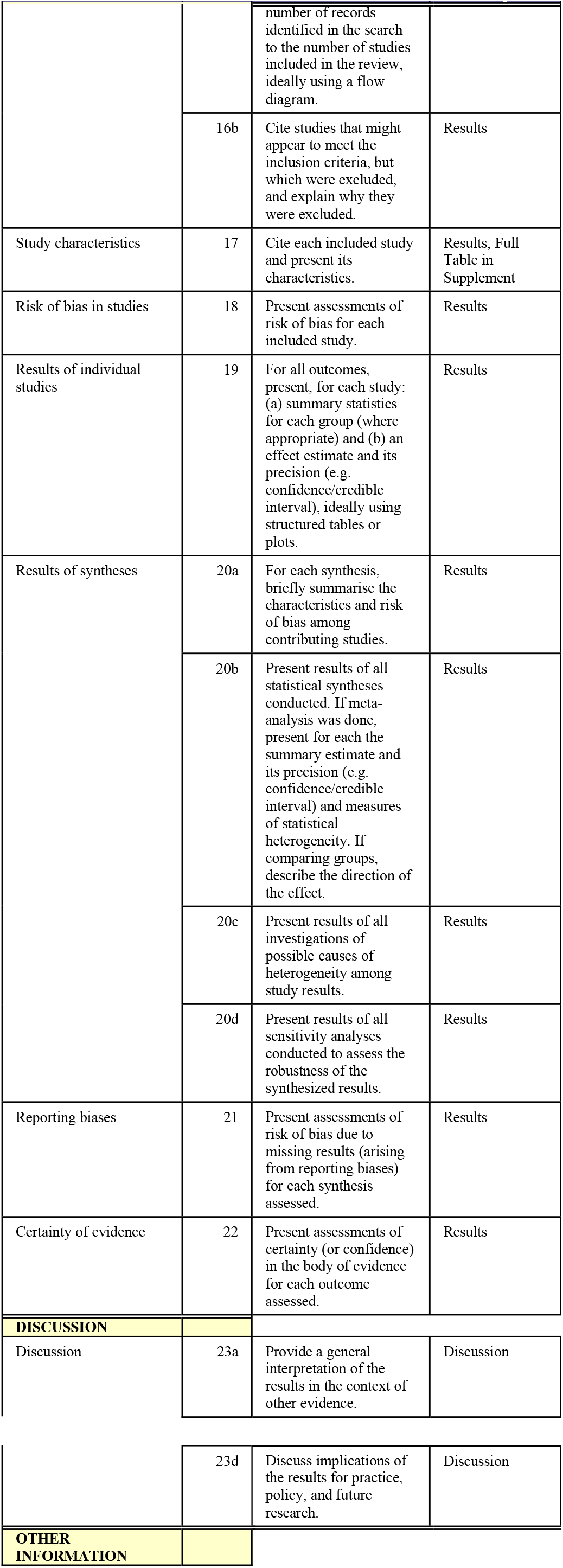
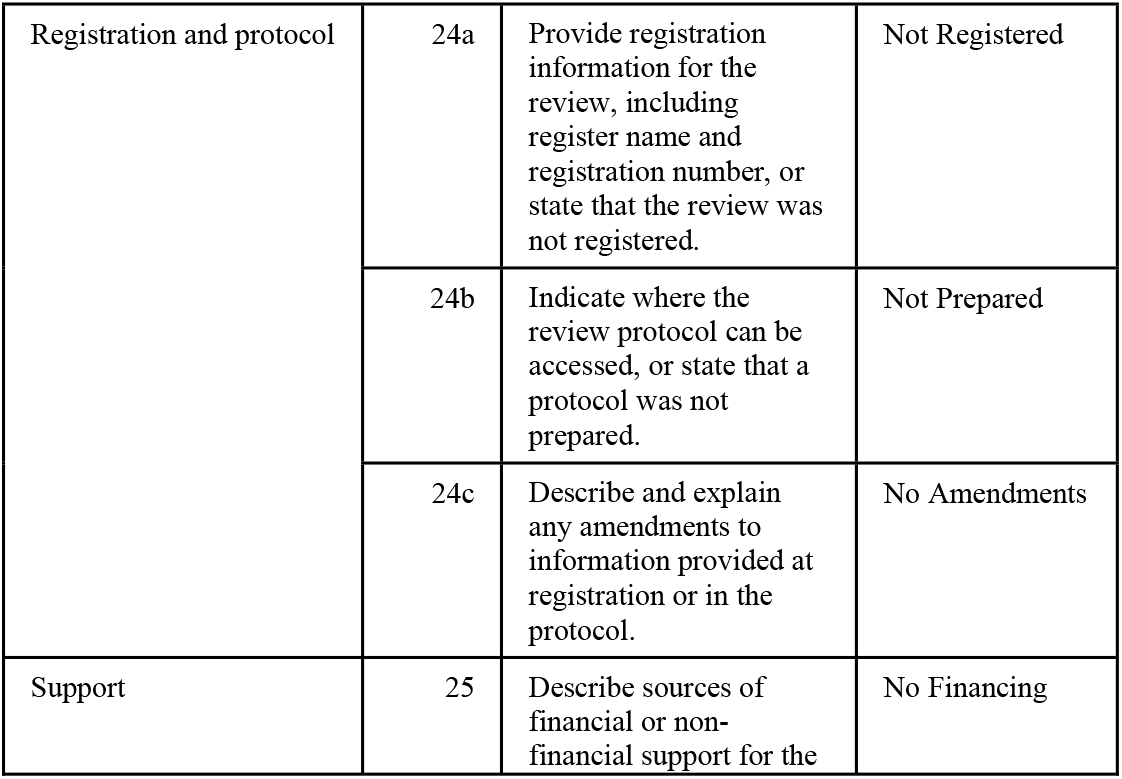
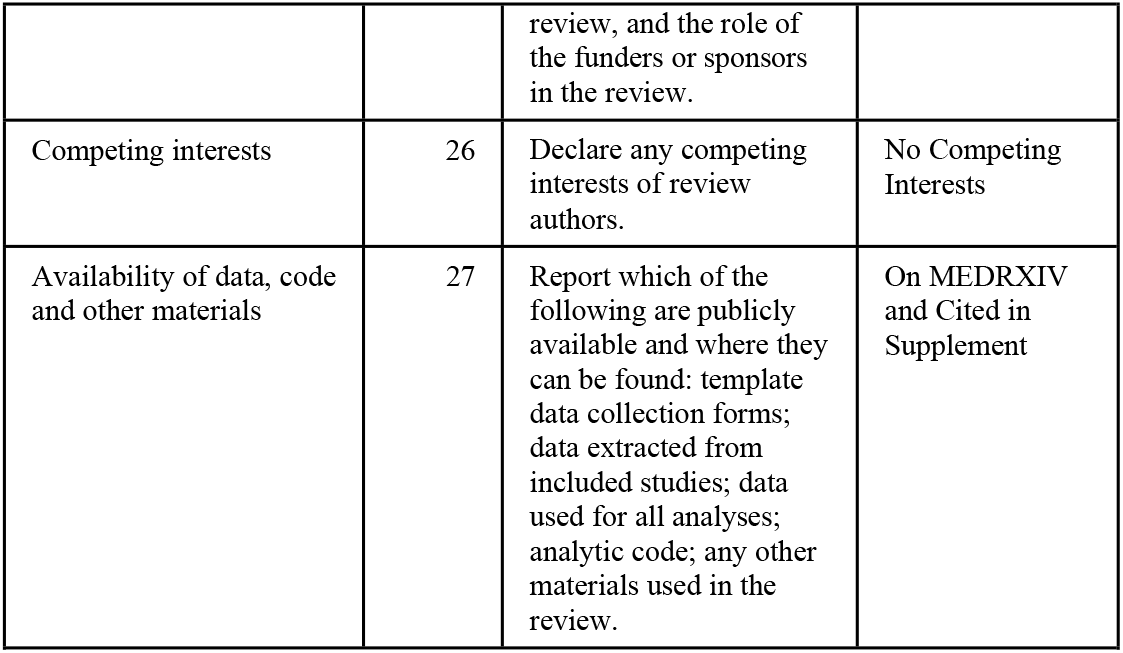
PRISMA Checklist.

